# Heg.IA: An intelligent system to support diagnosis of Covid-19 based on blood tests

**DOI:** 10.1101/2020.05.14.20102533

**Authors:** Valter Augusto de Freitas Barbosa, Juliana Carneiro Gomes, Maíra Araújo de Santana, Jeniffer Emidio de Almeida Albuquerque, Rodrigo Gomes de Souza, Ricardo Emmanuel de Souza, Wellington Pinheiro dos Santos

## Abstract

A new kind of coronavirus, the SARS-Cov2, started the biggest pandemic of the century. It has already killed more than 250,000 people. Because of this, it is necessary quick and precise diagnosis test. The current gold standard is the RT-PCR with DNA sequencing and identification, but its results takes too long to be available. Tests base on IgM/IgG antibodies have been used, but their sensitivity and specificity may be very low. Many studies have been demonstrating the Covid-19 impact in hematological parameters. This work proposes an intelligent system to support Covid-19 diagnosis based on blood testing. We tested several machine learning methods, and we achieved high classification performance: 95.159% *±* 0.693 of overall accuracy, kappa index of 0.903 *±* 0.014, sensitivity of 0.968 *±* 0.007, precision of 0.938 *±* 0.010 and specificity of 0.936 *±* 0.011. These results were achieved using classical and low computational cost classifiers, with Bayes Network being the best of them. In addition, only 24 blood tests were needed. This points to the possibility of a new rapid test with low cost. The desktop version of the system is fully functional and available for free use.

## 1. Introduction

A new kind of coronavirus started the biggest pandemic of the century. This virus received the name SARS-Cov2 to be related to severe acute respiratory syndrome coronavirus (Lin et al., 2020; Okba et al., 2020), one of the most dangerous consequences of Coronavirus Disease 19 (Covid-19). The clinical symptoms of Covid-19 patients include fever, cough, sore throat and some cases even gastrointestinal infection symptoms (Guo et al., 2020b). In the most severe cases it can lead to shortness of breath and death.

SARS-Cov2 probable origin is from a pangolin virus called Pangolin-CoV: they are 91.02% identical in whole-genome level (Zhang et al., 2020b). The first record of the SARS-Cov2 is from Wuhan, China in later December 2019. In May 2020 it have already spread to 215 countries in the world (WHO, 2020). The virus has a stronger human-to-human transmission capacity. Until May 2020 more than 3 people million were infected and more than 250 thousands of deaths (WHO, 2020). In this case, it is necessary quick and precise diagnosis tests (Beeching et al., 2020). The ground-truth test in Covid-19 diagnosis is the Reverse Transcription Polymerase Chain Reaction (RT-PCR) with DNA sequencing and identification (Döhla et al., 2020). However its result needs several hours to be ready (Döhla et al., 2020).

Tests based on IgM/IgG antibodies delivers results quickly, however they are nonspecific for Covid-19, and may have very low sensitivity and specificity (Patel et al., 2020; Burog et al., 2020; Egner et al., 2020; Döhla et al., 2020; Tang et al., 2020b). IgM/IgG tests do not detect the SARS-Cov2 presence directly, indeed they detect the serological evidence of recent infection. According to Li et al. (2020) positive response could be from of antibodies of other coronaviruses and flu viruses. The use of IgM/IgG rapid test kits as definitive diagnosis of Covid-19 in currently symptomatic patients is not recommended (Burog et al., 2020) because tests made in samples collected in the first week of illness have only 18.8% of sensitivity and 77.8% of specificity (Liu et al., 2020b). Döhla et al. (2020) compared the results of IgM/IgG with reverse transcription polymerase chain reaction (RT-PCR) in 59 patients and IgM/IgG tests obtained 36.4% of sensitivity and 88.9% of specificity.

IgG/IgM tests performed during the second week of the disease have 100% of sensitivity and 50% of specificity (Liu et al., 2020b). That means when the viral charge is high IgG/IgM tests reach high sensitivities and specificities. But in that cases the disease is in advanced levels (Guo et al., 2020a; Hoffman et al., 2020).

Many works have been demonstrating the Covid-19 impact in cardiovascular system and hematological characteristics (Fan et al., 2020; Tan et al., 2020; Gao et al., 2020; Liu et al., 2020a; Gunčar et al., 2018; Zheng et al., 2020). According to Turner et al. (2004) the coronaviruses, like SARS-Cov and SARS-Cov2, have as functional receptor the zinc metallopeptidase angiotensin-convertingenzyme 2 (ACE2), an enzyme presents in cell membranes of arteries, heart, lungs and other organs. ACE2 is implicated in heart function, hypertension and diabetes. Considering this fact, Zheng et al. (2020) pointed that Middle East respiratory syndrome-related coronavirus (MERS-CoV) and severe acute respiratory syndrome coronavirus (SARS-Cov) can cause acute myocarditis and heart failure. Some of the coronaviruses impacts in cardiovascular system are the increase in blood pressure, and increase in troponin I (hs-cTnI) levels (Zheng et al., 2020). Others works found that patients with Covid-19 can developed lymphopenia (low level of lymphocytes in the blood) (Fan et al., 2020; Tan et al., 2020; Liu et al., 2020a), leukopenia (few white blood cells). They can also have decrease in their hemoglobin levels, Absolute Lymphocyte Count (ALC) and Absolute Monocyte Count (AMC) (Fan et al., 2020). Patients that developed severe forms of the disease have significantly higher levels of hematological characteristics as Interleukin-6, D-dimer than patients that developed moderate form of the Covid-19 (Gao et al., 2020). Therefore, as Covid-19 is a disease that affects the blood, hematological exams can be used in diagnosis tests.

In the other hand, machine learning techniques have been used to diagnosis diseases by analyzing hematological parameters (Tanner et al., 2008; Luo et al., 2016; Gunčar et al., 2018). Machine learning is a subarea of the Artificial Intelligence in Computer Science. One of the mean goals of machine learning techniques is to perform pattern detection in databases.

In this way, it is necessary the development of one diagnosis support system that provide a quick result and with high sensibility even if the samples were be taken in early stages of the disease. In this context, blood tests utilization have some advantages. First, they are commonly used during medical screening. Besides that blood tests do not take too long to be ready and they have low price compared with other diagnosis methods.

In this work, we propose a diagnosis support system of the Covid-19 based in machine learning techniques. This pioneering system uses blood tests to diagnose Covid-19. We used the database provided by Hospital Israelita Albert Einstein located in São Paulo, Brazil. The database contains data from 5644 patients among them 559 patients were diagnosis with Covid-19 by using the gold standard method, the reverse transcription polymerase chain reaction (RTPCR) (Kaggle, 2020). More than one hundred laboratory exams are available in the database like blood counts and urine test. Thus, aiming to provide a quick diagnosis we performed the reduction of exams by choosing which tests are most relevant to the diagnosis. Firstly we made it by using attribute selection algorithms based on Evolutionary Search (ES) (Liang et al., 2000; Kim et al., 2000) and Particle Swarm Optimization (PSO) (Kennedy & Eberhart, 1995; Poli et al., 2007; Wang et al., 2007). In the following, we made a hand reduction aiming to obtain the lower test price, small period of time required to the exams to be ready and the lower amount of blood samples needed. Finally, we obtained a database with 24 blood tests. According to empirical information^1^, these exams may have turnaround time of 30 to 60 minutes in an emergency According to the Brazilian Society of Clinical Analysis, there is no reference document to establish standard turnaround times for the exams described above. context. So our proposal can be used as a rapid test.

We performed several experiments with different machine learning techniques as Multilayer Perceptron (Rosenblatt, 1958; Haykin, 2001), Support Vector Machine (Boser et al., 1992; Cortes & Vapnik, 1995), Random Trees, Random Forest (Breiman, 2001; Geurts et al., 2006), Bayesian networks and Naive Bayes (Cheng & Greiner, 2001; Haykin, 2001) and evaluated their results. Experiments show that Bayesian networks has superior performance with respect to other techniques. In order to choose this method, we considered six parameters: accuracy, sensibility, specificity, precision, recall, and kappa index.

This paper is organized as follows: Section 2 discuss some related works that found that Covid-19 can affect hematological characteristics. Section 3 presents the Kaggle database and reviews the theoretical machine learning methods concepts necessary to understand this work. It is also described how we manipulated Kaggle database and how we performed the experiments. Section 4 shows and analyzes the experimental results and the resulting desktop application we developed. We finalized the paper in section 6 with the conclusions.

## 2. Related works

Several studies have shown the importance of blood tests for diagnosis and indicative of the degree of severity of Covid-19. Fan et al. (2020) analyzed hematological indexes of 69 patients with Covid-19. All of them were treated at the National Center for Infectious Diseases (NCID), located in Singapore. Among these patients, 65 underwent complete blood count (CBC) on the day of admission. In addition, demographic information such as age, gender, ethnicity and location were made available for this research. 13.4% of patients needed intensive care unit (ICU) care, especially the elderly. During the first exams, 19 patients had leukopenia (few white blood cells) and 24 had lymphopenia (low level of lymphocytes in the blood), with 5 cases classified as severe (Absolute Lymphocyte Count (ALC) < 0.5 *×* 10^9^*/L*). The study also pointed out that patients who needed to be admitted to the ICU had lower ALC and a higher rate of Lactate Dehydrogenase (LDH). These data indicated that monitoring these parameters can help to identify patients who need assistance in the ICU. Also considering 9 patients who were in the ICU, the authors observed that the patients had a significant decrease in their hemoglobin levels, ALC and Absolute Monocyte Count (AMC) levels compared to the non-ICU group. ICU patients also tend to neuthophilia. The platelet count did not prove to be a factor for discrimination between the type of hospitalization.

Tan et al. (2020) also looked at complete blood count of patients (cured ones and also 12 patients who died from Covid-19). The authors sought to obtain key indicators of disease progression, as a way of supporting future clinical management decisions. In the case of patients who died, blood tests were monitored continuously throughout the treatment process. As in the previous study, the authors observed lymphopenia in this group. Based on this, the study then outlined a model (Time-LYM% model, TLM) for classifying disease severity and predicting prognosis. Thus, the blood lymphocyte percentage (LYM%) was divided into two cases, considering the first 10–12 days of symptoms: LYM% > 20% are classified as moderate cases and with a high chance of recovery. LYM% < 20% are classified as severe cases. In a second exam, 17–18 days after the first symptoms, patients with LYM% > 20% are recovering, patients with 5 <LYM% < 20% are in danger, and LYM% < 5% are in critical condition. In order to validate the model, the authors evaluated 90 patients with Covid-19. The consistency between Guideline and TLM-based disease classification was verified using kappa statistic (Kappa = 0.48). These results indicate that probably LYM% should be used together with other parameters for a better evaluation of Covid-19. Gao et al. (2020) observed hematological characteristics of 43 patients at Fuyang Second People’s Hospital. The patients had a diagnosis confirmed by the Covid-19 ground truth test, the fluorescent reverse transcription-polymerase chain reaction (RT-PCR). They were divided into two groups: the moderate group with 28 patients, and the severe group with 15 patients. The groups have no significant difference in age and sex. The blood tests observed were: Routine blood tests (white blood cell [WBC] count, lymphocyte count [LYM], mononuclear count [MONO], neutrophils count [NEU]) were performed on the blood samples. Blood biochemistry parameters (aspartate aminotransferase [AST], alanine aminotransferase [ALT], glucose [GLU], urea, creatinine [Cr], cystatin [Cys-c], uric acid [UA], and C-reactive protein [CRP]); Coagulation functions (the D-dimer [d-D], thrombin time [TT], prothrombin time [PT], fibrinogen [FIB], activated partial thromboplastin time [APTT]); rocalcitonin (PCT); and Interleukin-6 (IL-6). Using statistical tests, the study noted that the levels of GLU, CRP, IL-6, TT, FIB, and d-D were significantly higher in the severe group than in the mild group. Performing this analysis with ROC curves, the authors pointed out that the best indicators for predicting severity were IL-6 and d-D combined, with AUC of 0.840. The combination also achieved specificity of 96.4% and sensitivity of 93.3%, using tandem and parallel testing, respectively. These results indicate that patients with severe conditions would have abnormal coagulation.

Liu et al. (2020a) reported that lymphopenia and inflammatory cytokine storm are abnormalities commonly found in other infections caused by coronavirus, such as SARS-Cov and MERS-Cov. With that in mind, they studied 40 patients diagnosed with Covid-19 confirmed by throat-swab specimens analyzed with RT-PCR. The patients were treated at Wuhan Union Hospital in January, 2020. The information provided was: epidemiological, demographic, clinical manifestations and laboratory tests. Similar to the previous study, patients were divided into two groups: mild patients, with symptoms such as epidemiological history, fever or respiratory symptoms, and abnormalities in imaging tests; the second group with severe patients, patients should additionally have symptoms such as shortness of breath, oxygen saturation < 93%, respiratory > 30 times/min, or PaO_2_*/*FiO_2_ < 300 mmHg. 27 patients were classified in the first group, while 13 were classified in the second. The student reported that levels of fibrinogen, D-dimer, total bilirubin, aspartate transaminase, alanine transaminase, lactate dehydrogenase, creatine kinase, C-reactive protein (CRP), ferritin and serum amyloid A protein were significantly higher in the severe group. Futhermore, most severe patients presented lymphopenia, that can be related to the significantly decreased absolute counts of T cells, especially CD8+ T cells, while white blood cells and neutrophils counts were higher.

These studies have pointed out that hematological parameters can be indicators of the risk factors and degree of severity of Covid-19. The identification of these parameters can be essential to facilitate clinical care for each group of patients. In this sense, the development of intelligent systems based on blood tests is useful. Faced with the pandemic scenario, in which most hospitals are full, decision support systems can facilitate clinical management. Thus, it can increase the assertiveness in the treatment for each case and, consequently, the number of lives saved.

Gunčar et al. (2018) proposed a system based on machine learning for analyzing blood tests and predicting hematological diseases. The models were developed with data from the University Medical Center of Ljubljana, which were collected between the years 2005 and 2015. In this case, 43 diseases and 181 parameters or attributes were selected to generate a first model (SBAHEM181). In addition to it, a second model with 61 parameters was also developed (SBA-HEM061). The selection of the smaller group was made according to the frequency of use. Regarding the missing values (about 75%), the authors filled in with median values for each attribute. For the development of the intelligent system, classic classifiers such as Support Vector Machines, Naive Bayes and Random Forest were tested, with the latter being chosen. The simulations were repeated 10 times using 10-fold cross validation. Finally, the models SBA-HEM181 and SBA-HEM061 reached an accuracy of 57% considering all the diseases chosen. By restricting the prediction to five classes, the systems achieved an accuracy of 88% and 86%, respectively. These results pointed to the possibility of detecting diseases through blood tests using classic classifiers.

## 3. Materials and methods

In this work we explore, evaluate and analyze the performance of various machine learning techniques to predict Covid-19 diagnosis based on different clinical exams configurations. In addition, we also propose an intelligent system in a desktop application format.

### 3.1. Proposed method

In this work we propose a support system for Covid-19 detection using clinics blood exams called Heg.IA. Heg.IA is a smart system pioneered in the rapid detection of Covid-19 through blood tests. The basic operation of this system is: in emergency situations, the medical team should order some blood exams of patients that have characteristic symptoms of Covid-19; with the results in hands the healthcare professional could insert them in a desktop system. In the following, the system will inform whether the patient is infected with Covid-19 or not. The result will be presented with the respectively classification probabilities, helping physicians to making their final decision. Figure 1 shows a general schematic of the solution.

**Figure 1:**
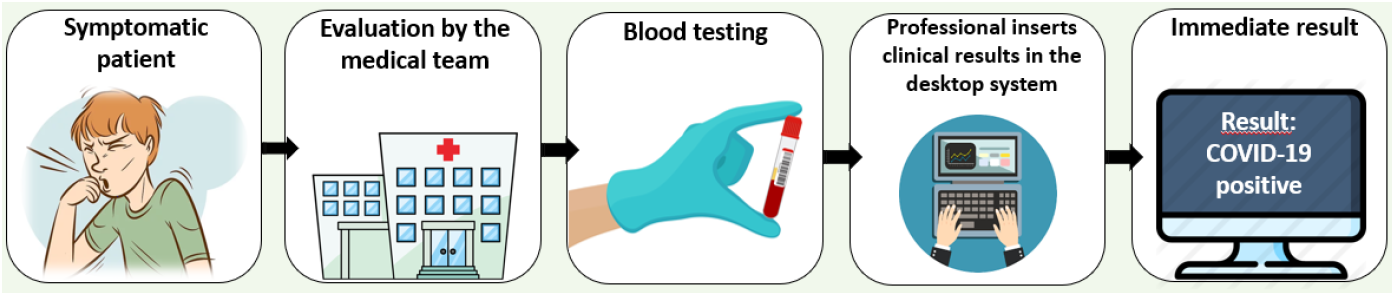
General proposal to diagnostic support system for the detection of Covid-19. Patients with symptoms of Covid-19 should go to health centers. The medical team must ask blood tests. Then, the results can be entered into the Heg.IA system, which will indicate if the patients is infected with SARS-Cov2.

### 3.2. Database

For the development of this project, we used the database provided by Hospital Israelita Albert Einstein located in São Paulo, Brazil. The database was made available in the competition format (Kaggle, 2020). The database contains the results of more than one hundred laboratory exams, such as blood count, tests for the presence of viruses such as influenza A, and urine tests, of 5644 patients. Among these patients, 559 of them are infected with SARS-Cov2. The Covid-19 diagnosis were made with reverse transcription polymerase chain reaction (RT-PCR) with DNA sequencing and using additional laboratory tests during a visit to the hospital (Kaggle, 2020). The clinical data provided in the data were normalized to have mean of zero and standard deviation one (Kaggle, 2020).

### 3.3. Database Pre-processing

In order for the data provided in the database to be used as input parameters for a machine learning method, it was necessary to assign integer numeric values to the columns containing categorical data. These columns have results of exams for detecting whether a pathogen is present or not or the appearance of patient’s urine, for example. Besides that, considering that 90,1% of data is from healthy patients it is reasonable to assume that missing values are within the normal range.

In this way, for the parameters about the presence of pathogens it was assigned the value zero (0) for pathogens’ absence and one (1) for pathogens’ presence. Thus, for the attribute with SARS-Cov2 exam result, which gives the information whether the patient has Covid-19 or not, it was assigned the value 0 for cells marked as negative and 1 for cells with positive result. Following this procedure, the attributes Respiratory Syncytial Virus, Influenza A, Influenza B, Parainfluenza 1, Coronavirus NL63, Rhinovirus/ Enterovirus, Mycoplasma pneumoniae, Coronavirus HKU1, Parainfluenza 3; Chlamydophila pneumoniae, Adenovirus, Parainfluenza 4, Coronavirus 229E, Coronavirus OC43, Influenza A H1N1 2009, Bordetella pertussis, Metapneumovirus, Parainfluenza 2, Influenza B evaluated with rapid test, Influenza A evaluated with rapid test, and Strepto A, it was assigned the value 0 for the not detected cases and 1 for abnormalities detected, and the value 2 was used for missing values.

The changes made to the attributes pertaining to urine tests followed the same pattern of the pathological examinations. Thus, for the attributes Urine-Esterase, Urine-Hemoglobin, Urine-Bile pigments, Urine-Ketone Bodies, Urine-Protein, Urine-Hyaline cylinders, Urine-Granular cylinders and Urine-Yeasts it was put on 0 for absent cases and missing values and the value 1 for the presence. In Urine Aspect column it was adopted 0 for clear and missing values, 1 for lightly cloudy class and 2 for cloudy class. In Urine-Crystals the absense category and missing values were replaced by 0, Amorphous Urate replaced by 1, Calcium Oxalate replaced by 2. The light yellow category and missing values from Urine-Color was assigned as 0, yellow as 1 and orange as 2. Whereas “Urine – Urobilinogen the results classified as normal were replaced by 1, not done or missing value by 0. Lastly the values in Urine – Leukocytes less than 10000 were replaced by 0 and the values greater or equal than 10000 by 1. Considering that the database is normalized with mean of zero the missing values from columns containing numeric values were filled by 0. In other words, the missing values were filled with the mean. This is due to our presumption that most patients in the database set are healthy. So we assumed that the mean is within in normal range.

Besides that, the low number of patients with Covid-19 in the database might bias the classification to the majority class (healthy patients class). In view of that, we performed a synthetic balancing by using Synthetic Minority Over-sampling TEchnique (SMOTE). It is an oversampling technique for generating synthetic samples from the minority class. SMOTE uses linear combinations of two similar samples to construct new data (Blagus & Lusa, 2013).

After changing categorical data and handle with missing values it was possible to build in up a database used to predict Covid-19 diagnosis.

### 3.4. Attribute Selection

The database constructed during the pre-processing was submitted to feature selection using Particle Swarm Optimization (PSO) (Kennedy & Eberhart, 1995; Poli et al., 2007; Wang et al., 2007) and Evolutionary Search (ES) (Liang et al., 2000; Kim et al., 2000). Both using 20 individuals and 500 iterations. The goal of the attribute selection is to find which exams are more relevant for classification tasks and also to reduce the number of required exams for diagnostic support and consequently its price. Thus the solution can be extend to public healthcare center with low costs.

We chose to use PSO and ES algorithms, since they are well established methods for feature selection. Both feature selection methods are widely used due to their global search ability. PSO algorithm uses a population of randomly generated particles. In this approach, each particle corresponds to randomly generated solution and have an associated velocity and position. ES, on the other hand, is a version of genetic algorithms, which emulates Darwinism principle of natural selection and genetic reproduction. Thus, this method uses the evolution principles to assess the best solution to a given problem.

The feature selection implementation resulted three databases in total: The original database with 108 attributes: SARS-Cov2 (PSO) and SARS-Cov2 (ES) with 63 and 62 attributes, respectively. The acronyms PSO and ES in database’ names indicate that the database was obtained by attribute selection using PSO and ES respectively.

After selecting attributes with PSO and ES, a last selection was made manually. This aims to reduce the number of tests required for the diagnosis and speeding up the process. Consequently, we have also achieved a reduction in exam costs. The selection was made based on the prices of each of the listed exams. They were removed one by one, and the performance of the classification was evaluated at each step. Thus, it was possible to significantly decrease the number of attributes, with minor impacts in the performance of the system. Finally, a fourth database was generated with only 24 attributes. The base was named SARS-CoV2 (CheapExams). This complete process is described in the diagram in Figure 2.

**Figure 2:**
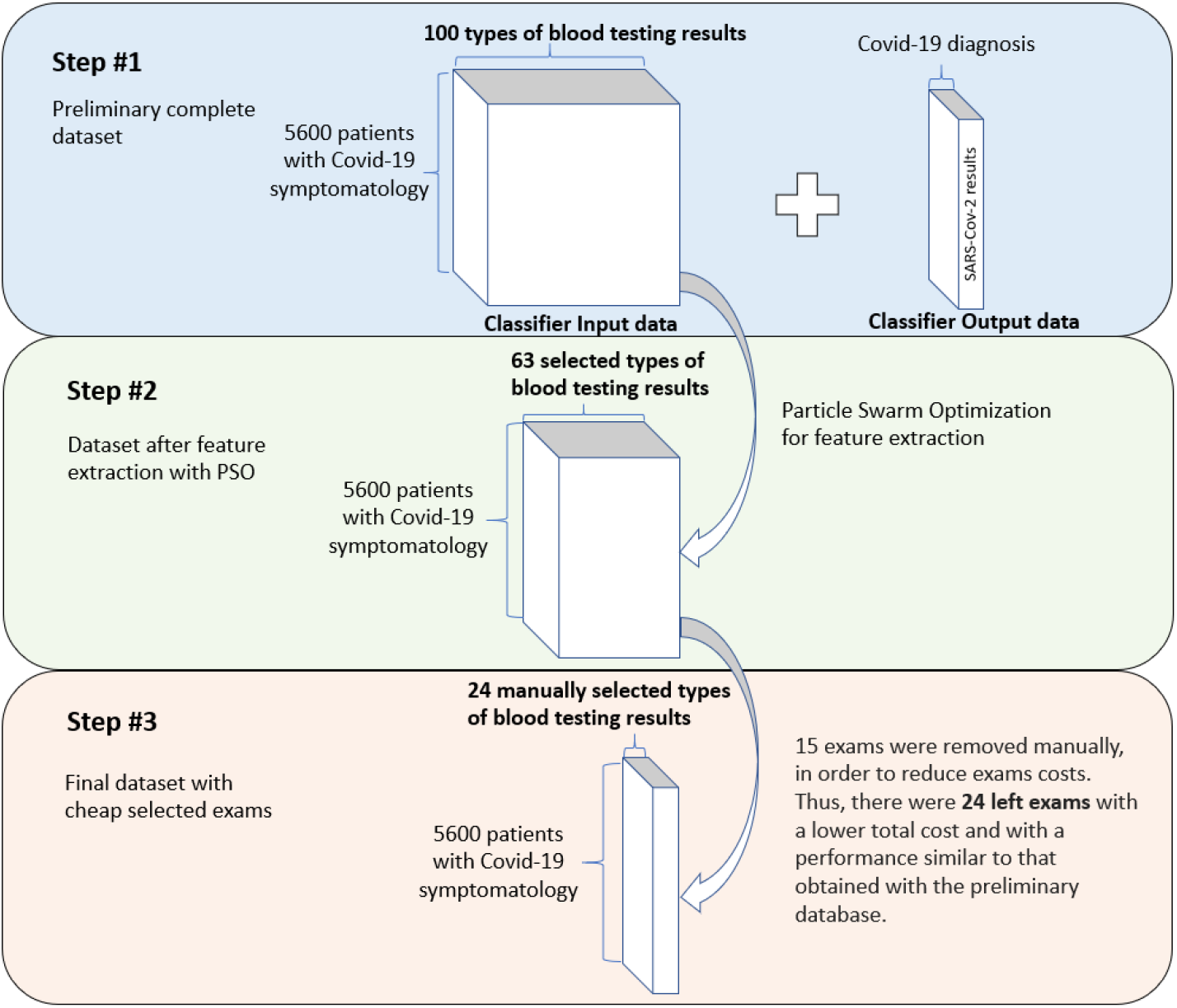
Description of feature extraction process: PSO and ES algorithms were applied to the original database, resulting in 63 attributes. In the following, 24 attributes were selected manually with a lower total cost and with a performance similar to the preliminary database.

### 3.5. Classification

#### 3.5.1. Multilayer Perceptron

In 1958, Frank Rosenblatt (Rosenblatt, 1958) proposed the perceptron model for supervised learning. Rosenblatt was psychologist and one of the pioneers in the concept of artificial neural networks. His perceptron model is the simplest form of neural network used for binary classifications of linearly separable patterns. It consists of a single neuron with adjustable synaptic weights and a bias (Haykin, 2001).

Multilayer Perceptron (MLP) is a generalization of the single layer perceptron. It consists of a feed-foward network with an input layer, hidden layers and one output layer. The addition of hidden layers allows to the network the ability to classify more complex problems than single layer perceptron such as image classification (Lerner et al., 1994; Phung et al., 2005; Barbosa et al., 2020).

MLP training is performed in a supervised way, through the error correction learning rule. This rule adjusts the synaptic weights in a way that the network output becomes closer to the expected output. This method usually uses an error backpropagation algorithm to adjust these weights. Based on a gradient descent, backpropagation proceeds in two phases: propagation and backpropagation. In the propagation phase, an output is obtained for a given input pattern. In backpropagation phase, an error is calculate using the desired output and the output obtained in the previous phase. Then, the error is used to update the connection weights. Thus backpropagation aims to iteratively minimize the error between the network output obtained and the desired output (Haykin, 2001).

MLPs and other artificial neural networks architectures like Extreme Learning Machines have been commonly used in support diagnosis applications, e.g. breast cancer diagnosis over breast thermography (de Vasconcelos et al., 2018; Pereira et al., 2020b; Santana et al., 2020; Pereira et al., 2020a,c; Santana et al., 2018; Rodrigues et al., 2019), mammography images (de Lima et al., 2016; Lima et al., 2015; de Lima et al., 2014; Silva et al., 2020; Cordeiro et al., 2017, 2016; de Lima et al., 2014; Cruz et al., 2018), and multiple sclerosis diagnosis support (Commowick et al., 2018).

#### 3.5.2. Support Vector Machine

Created by Vladimir Vapnik and Alexey Chervonenkis (Boser et al., 1992; Cortes & Vapnik, 1995), in 1963, the support vector machine (SVM) perform a nonlinear mapping on the database in a space of high dimension called feature space. So the algorithm builds a linear decision surface, called hyperplane, to separate distinct classes (Cortes & Vapnik, 1995). SVMs have been widely used in several medical applications, e.g. breast cancer diagnosis over breast thermography (de Vasconcelos et al., 2018; Pereira et al., 2020b; Santana et al., 2020) and mammography images (de Lima et al., 2016; Lima et al., 2015; de Lima et al., 2014; Silva et al., 2020; Cordeiro et al., 2017, 2016; de Lima et al., 2014; Cruz et al., 2018).

Thus, the training process of a support vector machine aims to find the hyperplane equation which maximizes the distance between it and the nearest data point. That hyperplane is called optimal hyperplane (Haykin, 2001). This machine uses support vector learning, which is a subset of the training data. SVM algorithm is well known for its ability to provide good generalization performance (Haykin, 2001). However, this performance may decrease by increasing the complexity of the hyperplane. The type of the machine varies with the type of kernel used to build the optimal hyperplane. Table 1 shows the kernel functions used in the present study.

**Table 1:** Kernel functions of SVM.

| Type of SVM | Kernel function |
| --- | --- |
| Polynomial | $K(\mathbf{x}, \mathbf{y}) = (\mathbf{x} \bullet \mathbf{y} + 1)^E$ |
| Radial Basis Function (RBF) | $K(\mathbf{x}, \mathbf{y}) = \exp(-\gamma (\mathbf{x} - \mathbf{y}) \bullet (\mathbf{x} - \mathbf{y}))$ |

#### 3.5.3. Decision Trees

Decision trees are a type of supervised machine learning model. They can used in classification and regression problems. In most cases, trees have nodes responsible to store information. Essentially, in a tree there are four types of nodes: root, leaf, parent and child. The starting point is the Root node that has the highest hierarchical level. One node may connect to another, establishing a parent-child relationship, in which a parent node generate a child node. The terminal nodes is composed by Leaf nodes. Thus they have no children, and represent a decision. In this fashion, using such trees, the algorithm makes a decision after following a path that starts from the root node and reaches a leaf node. There are several types of decision trees. They usually differ from the way the method goes through the tree structure. Random Tree and Random Forest methods are the most main ones.

According to Geurts et al. (2006) Random Tree algorithm uses a tree built by a stochastic process. This method considers only a few randomly selected features in each node of the tree.

In the other hand, Random Forest algorithm, consists in a collection of trees. These trees hierarchically divide the data, in other that, each tree votes for a class of the problem. At the end, the algorithm choose the most voted class as the prediction of the classifier Breiman (2001).

#### 3.5.4. Bayesian Network and Naive Bayes

Bayes Net and Naive Bayes are classifiers based on Bayes’ Decision Theory. They are also called the test procedure by the Bayes hypothesis. Bayesian classifiers seek to find a minimum mean risk. By considering a set of correct decisions and a set of incorrect decisions, they use conditional probability to create the data model. The product of the frequency of each decision with the cost involved in making the decision results in the weights (Haykin, 2001). Bayesian networks behave like a linear classifier for a Gaussian distribution. Its behavior is comparable to that of a single layer perceptron.

In the standard Bayes Network algorithm, it assesses the probability of occurrence of a class from the values given by the others. Thus, this method assumes dependence between the features. Naive Bayes, in turn, considers that all features are independent of each other, being only connected to the class (Cheng & Greiner, 2001). This method does not allow dependency between features. Considering this assumption represents an unrealistic condition, the algorithm is considered “naive”.

### 3.6. Parameters settings of the classifiers

The experiments were made by using the following techniques: SVM with polynomial kernel of degree (*E*) 1, 2, and 3 and RBF kernel with *γ* of 0.01; MLP with 50 and 100 neurons in hidden layer; Random Forest with 10, 20, 30, *…*, 100 trees; Random Tree; Naive Bayes; Bayesian Network. As a evaluate method we choose to perform a 10-fold cross validation and each configuration was executed 25 times.

### 3.7. Metrics

We chose six metrics to evaluate the performance of diagnostic tests: accuracy, precision, sensitivity, specificity, recall, and precision. Accuracy is the probability that the test will provide correct results, that is, be positive in sick patients and negative in healthy patients. In other words, it is the probability of the true positives and true negatives among all the results. The recall and sensitivity metrics can be calculated mathematically in the same way. They are the rate of true positives, and indicate the classifier ability to detect correctly people with Covid-19. However, they are commonly used in different contexts. In machine learning context, the term Recall is common. However, in the medical world, the use of the sensitivity metric is more frequent. Precision, on the other hand, is the fraction of the positive predictions that are actually positive. Specificity is the capacity of classifying healthy patients as negatives. It is the rate of true negatives. Finally, the Kappa index is a very good measure that can handle very well both multi-class and imbalanced class problems, as the one proposed here. These six metrics allow to discriminate between the target condition and health, in addition to quantifying the diagnostic exactitude (Borges, 2016). They can be calculated according to the equations in Table 2.

**Table 2:** Metrics used to evaluate classifiers performance: overall accuracy, sensitivity (recall), precision, specificity, and kappa index.

| Metrics | Equation |
| --- | --- |
| Accuracy | $Accuracy = \frac{TP + TN}{TP + TN + FP + FN}$ |
| Sensitivity and Recall | $Sensitivity = Recall = \frac{TP}{TP + FN}$ |
| Precision | $Precision = \frac{TP}{TP + FP}$ |
| Specificity | $Specificity = \frac{TN}{TN + FP}$ |
| Kappa index | $k = \frac{\rho_o - \rho_e}{1 - \rho_e}$ |

In Table 2, TP is the true positives, TN is the true negatives, FP is the false positives, and FN the false negatives, *ρ_o_* is observed agreement, or accuracy, and *ρ_e_* is the expected agreement, defined as following:

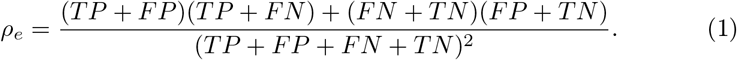

## 4. Results

### 4.1. Feature Selection

During the features selection procedure, we first performed selections based on PSO and ES algorithms. The features selected by both methods resulted in equivalent classification performances, as described in Table 3. Considering the values of mean and standard deviation of the six metrics, the two methods presented similar performance: Mean accuracy of 99.0865, mean kappa index of 0.9817, sensitivity of 0.9938, mean precision of 0.9880, and mean specificity of 0.9880. In addition, most selected exams were the same. Thus, we chose to select manually the cheaper exams from the ones selected by PSO algorithm. Figure 3 details the list of exams for each situation. In the first list of Figure 3 (in blue) are all 108 exams from the original database. The second (in green) shows the 63 most relevant exams selected by PSO algorithm. In the third list, we present the final set of 24 exams.

**Table 3:**
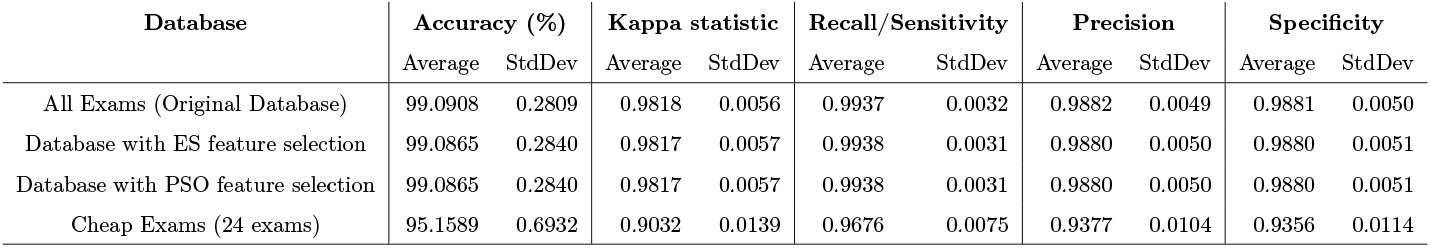
Comparison of classification performance using Bayes Net among the three databases: With all 107 attributes (original database), with attribute extraction using PSO, and with the 24 cheap exams. The results show that the metrics had a minimal reduction with the decrease of the attributes. exams.

**Figure 3:**
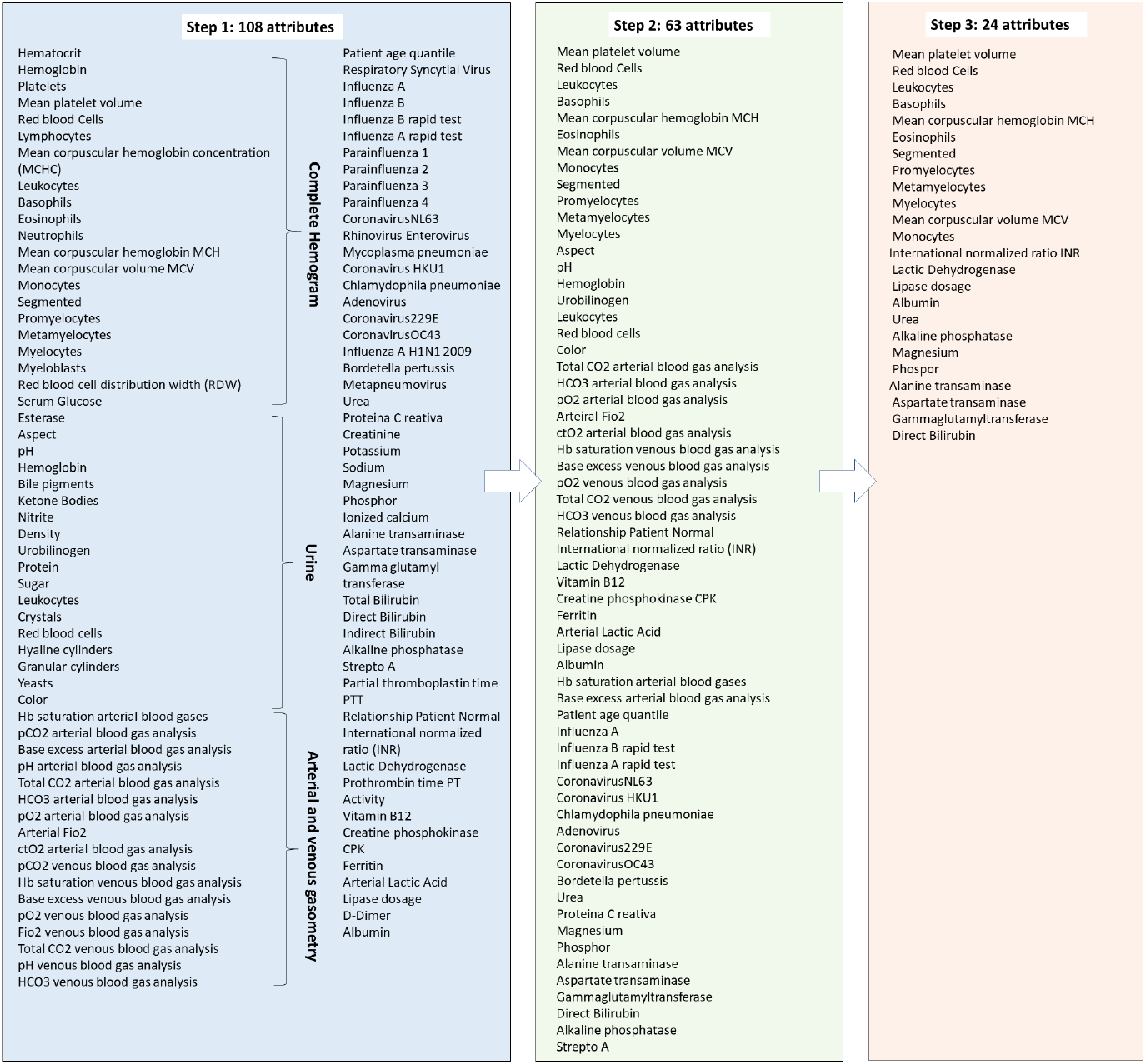
Description of the attributes in each of the bases used. Initially, the original database had 108 exams or attributes. After applying the PSO method for extracting attributes, a second base was formed with 63 attributes. Finally, 24 exams were selected manually

### 4.2. Classification

This section presents the results obtained in the classification phase. Tables 4 and 5 show the results of mean and standard deviation for all studied classifiers. Table 4 presents the results for the database with the complete set of exams. Table 5 shows the results for the reduced database, with 24 cheaper exams.

**Table 4:** Results of experiments with multiple classifiers using the original database (All exams). pandemic.

|  |  | SARS-Cov2 (All exams) |  |  |  |
| --- | --- | --- | --- | --- | --- |
| Classifier |  | Kappa | Accuracy | Recall | Precision |
| SVM Kernel<br>Polynomial E1 | Mean | 0.930 | 96.460 | 0.960 | 0.960 |
|  | Std Dev | 0.010 | 0.570 | 0.010 | 0.010 |
| SVM Kernel<br>Polynomial E2 | Mean | 0.930 | 96.330 | 0.960 | 0.960 |
|  | Std Dev | 0.010 | 0.580 | 0.010 | 0.010 |
| SVM Kernel<br>Polynomial E3 | Mean | 0.920 | 96.080 | 0.960 | 0.960 |
|  | Std Dev | 0.010 | 0.580 | 0.010 | 0.010 |
| SVM Kernel RBF | Mean | 0.910 | 95.630 | 0.960 | 0.960 |
|  | Std Dev | 0.010 | 0.620 | 0.010 | 0.010 |
| MLP (50 neurons in<br>the hidden layer) | Mean | 0.900 | 95.060 | 0.950 | 0.950 |
|  | Std Dev | 0.020 | 0.840 | 0.010 | 0.010 |
| MLP (100 neurons in<br>the hidden layer) | Mean | 0.900 | 95.060 | 0.950 | 0.950 |
|  | Std Dev | 0.020 | 0.840 | 0.010 | 0.010 |
| Random Forest (10<br>iterations) | Mean | 0.950 | 97.350 | 0.970 | 0.970 |
|  | Std Dev | 0.010 | 0.500 | 0.010 | 0.000 |
| Random Forest (20<br>iterations) | Mean | 0.960 | 97.900 | 0.980 | 0.980 |
|  | Std Dev | 0.010 | 0.430 | 0.000 | 0.000 |
| Random Forest (30<br>iterations) | Mean | 0.960 | 98.060 | 0.980 | 0.980 |
|  | Std Dev | 0.010 | 0.420 | 0.000 | 0.000 |
| Random Forest (40<br>iterations) | Mean | 0.960 | 98.130 | 0.980 | 0.980 |
|  | Std Dev | 0.010 | 0.430 | 0.000 | 0.000 |
| Random Forest (50<br>iterations) | Mean | 0.960 | 98.160 | 0.980 | 0.980 |
|  | Std Dev | 0.010 | 0.430 | 0.000 | 0.000 |
| Random Forest (60<br>iterations) | Mean | 0.960 | 98.190 | 0.980 | 0.980 |
|  | Std Dev | 0.010 | 0.430 | 0.000 | 0.000 |
| Random Forest (70<br>iterations) | Mean | 0.960 | 98.200 | 0.980 | 0.980 |
|  | Std Dev | 0.010 | 0.440 | 0.000 | 0.000 |
| Random Forest (80<br>iterations) | Mean | 0.960 | 98.220 | 0.980 | 0.980 |
|  | Std Dev | 0.010 | 0.440 | 0.000 | 0.000 |
| Random Forest (90<br>iterations) | Mean | 0.960 | 98.230 | 0.980 | 0.980 |
|  | Std Dev | 0.010 | 0.440 | 0.000 | 0.000 |
| Random Forest (100<br>iterations) | Mean | 0.960 | 98.240 | 0.980 | 0.980 |
|  | Std Dev | 0.010 | 0.450 | 0.000 | 0.000 |
| Bayesian Network | Mean | **0.980** | **99.090** | **0.990** | **0.990** |
|  | Std Dev | **0.010** | **0.270** | **0.000** | **0.000** |
| Naïve Naves | Mean | 0.950 | 97.420 | 0.970 | 0.980 |
|  | Std Dev | 0.010 | 0.470 | 0.000 | 0.000 |
| Random Tree | Mean | 0.840 | 91.800 | 0.920 | 0.920 |
|  | Std Dev | 0.030 | 1.600 | 0.020 | 0.020 |

**Table 5:**
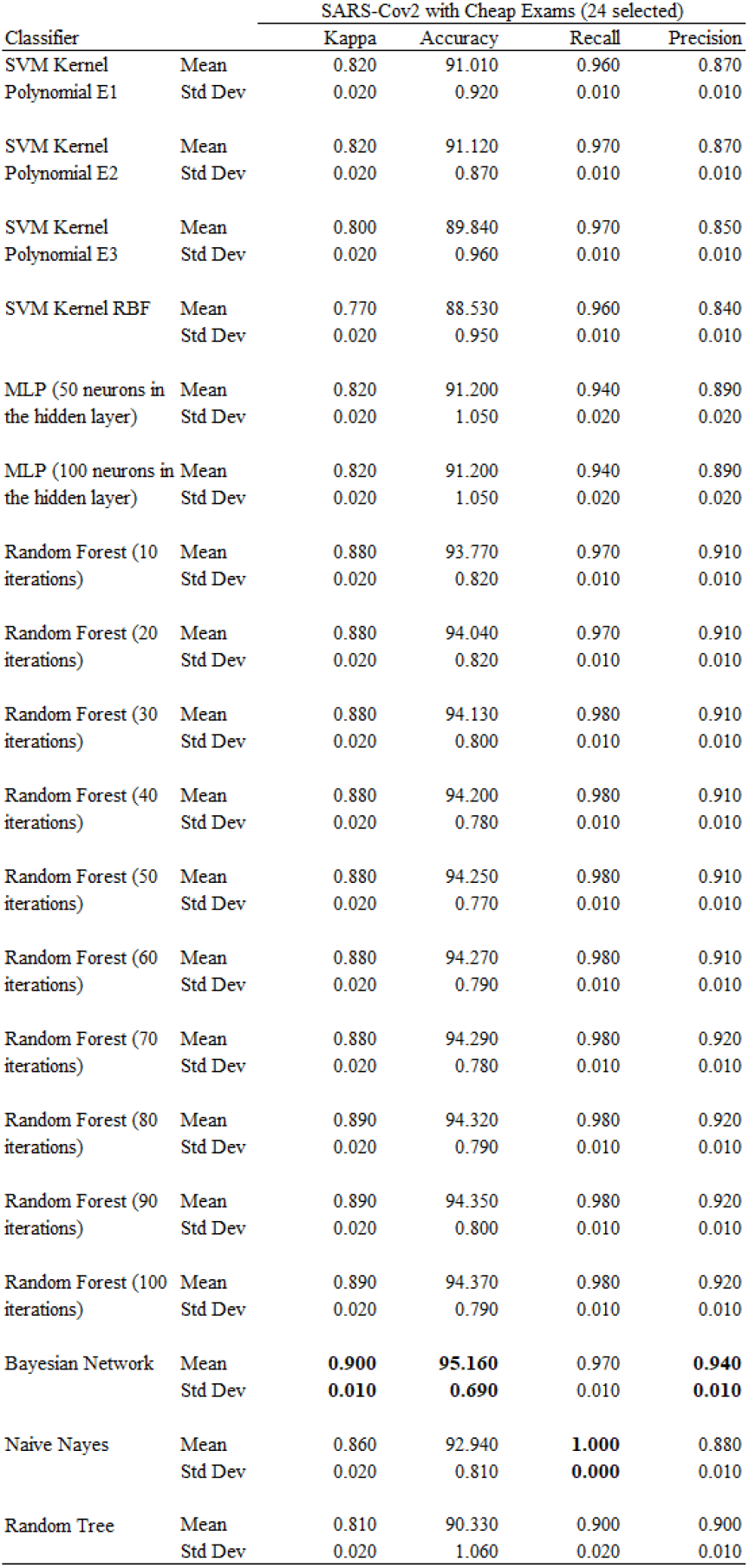
Results of experiments with multiple classifiers using the database with 24 attributes (Cheap exams).

Figures 4 and 5 show the performance of the different methods for diagnosing SARS-Cov2 using only the 24 cheapest exams. Figure 4 presents the results of accuracy of these methods, while Figure 5 shows the results of kappa index. Both graphs contain statistical information from the 25 repetitions performed for each configuration. As previously mentioned, 10 folds cross-validation was used in all tests.

**Figure 4:**
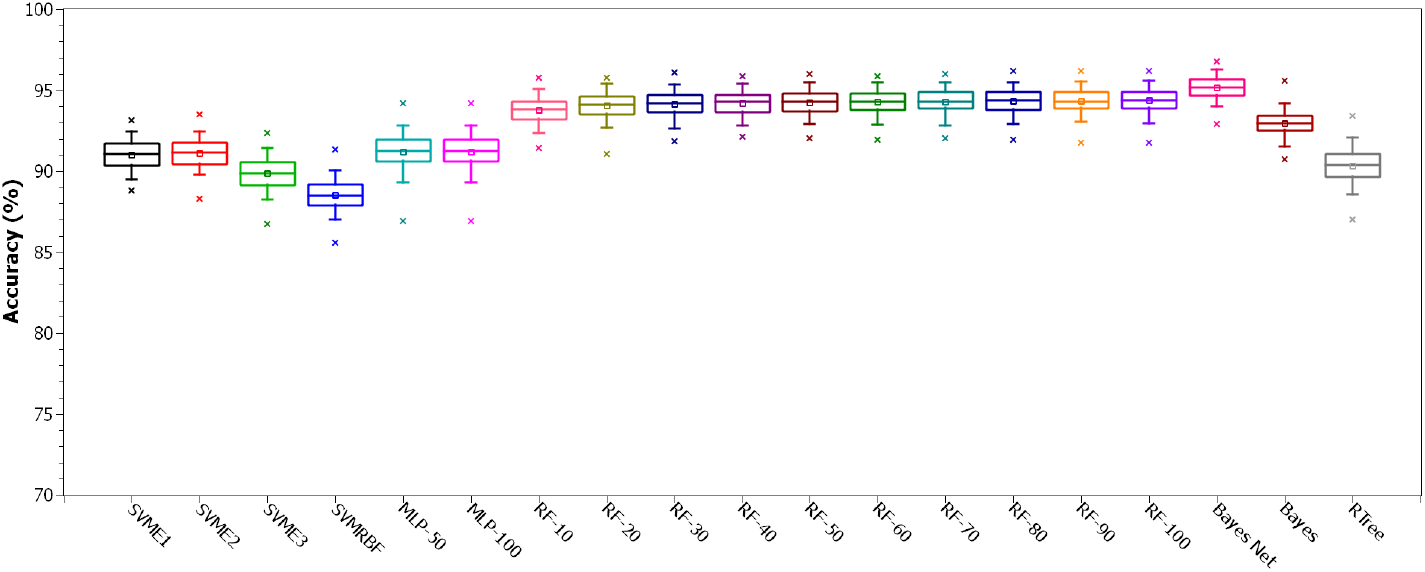
Comparison of Accuracy between configurations for Cheap Exams database.

**Figure 5:**
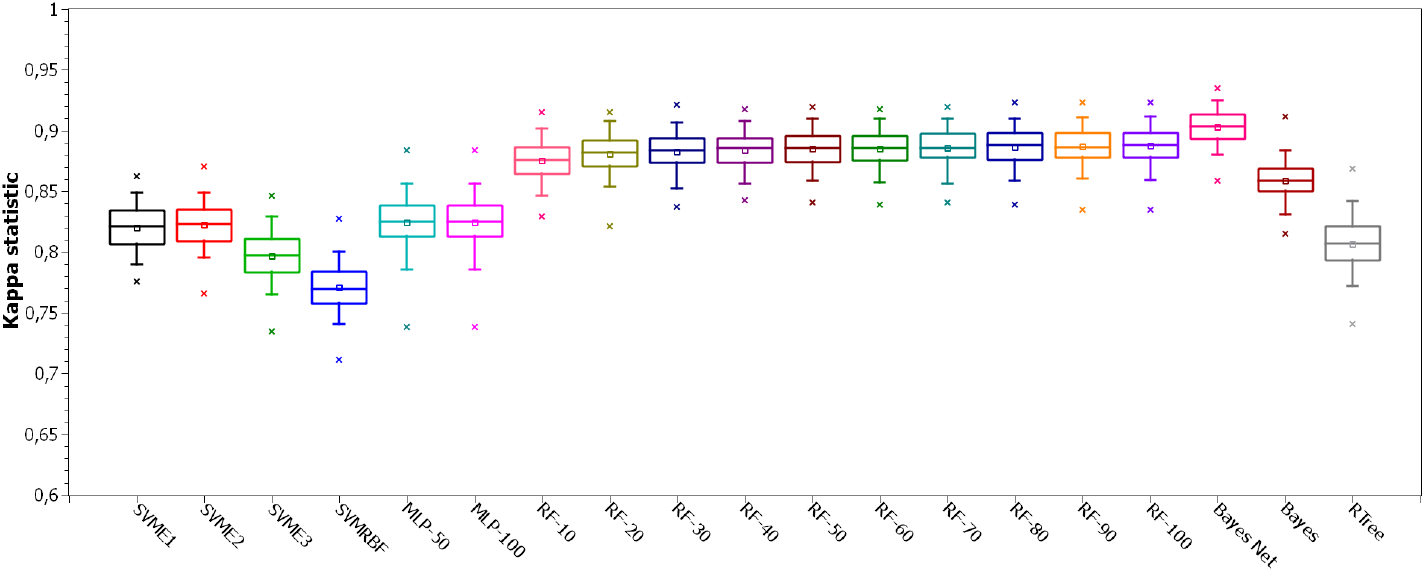
Comparison of Kappa statistic between configurations for Cheap Exams database.

From the results in Figures 4 and 5 it is possible to see that Bayes Net method overcame the others. Therefore, in Table 3, we compare the results of this method to the complete database and the two databases with reduced amount of exams.

### 4.3. HegIA Desktop Application

The prototype of the developed system is fully functional in a desktop version. It works like this: The health professional will be able to type the test results in the application. In this first version of the system, the exams must be entered in the unit indicated on the screens. Figures 6 and 7 show the list of exams with their respective fields. Figure 6 shows general exams, while Figure 7 exams belonging to the blood count. After completing the exams, the professional can select the “predict” option. Although the system works more efficiently with all fields filled in, it will also provide prediction with missing esams. Finally, it is possible to view the screen in Figure 8, where the positive or negative diagnosis for Covid-19 is presented. In addition to the diagnosis, the screen presents the values of the main classification metrics, helping the professional in his decision making. On the screen it is also possible to view results to predict hospitalization. This information is nearing completion, and the full system will be presented in a near future.

**Figure 6:**
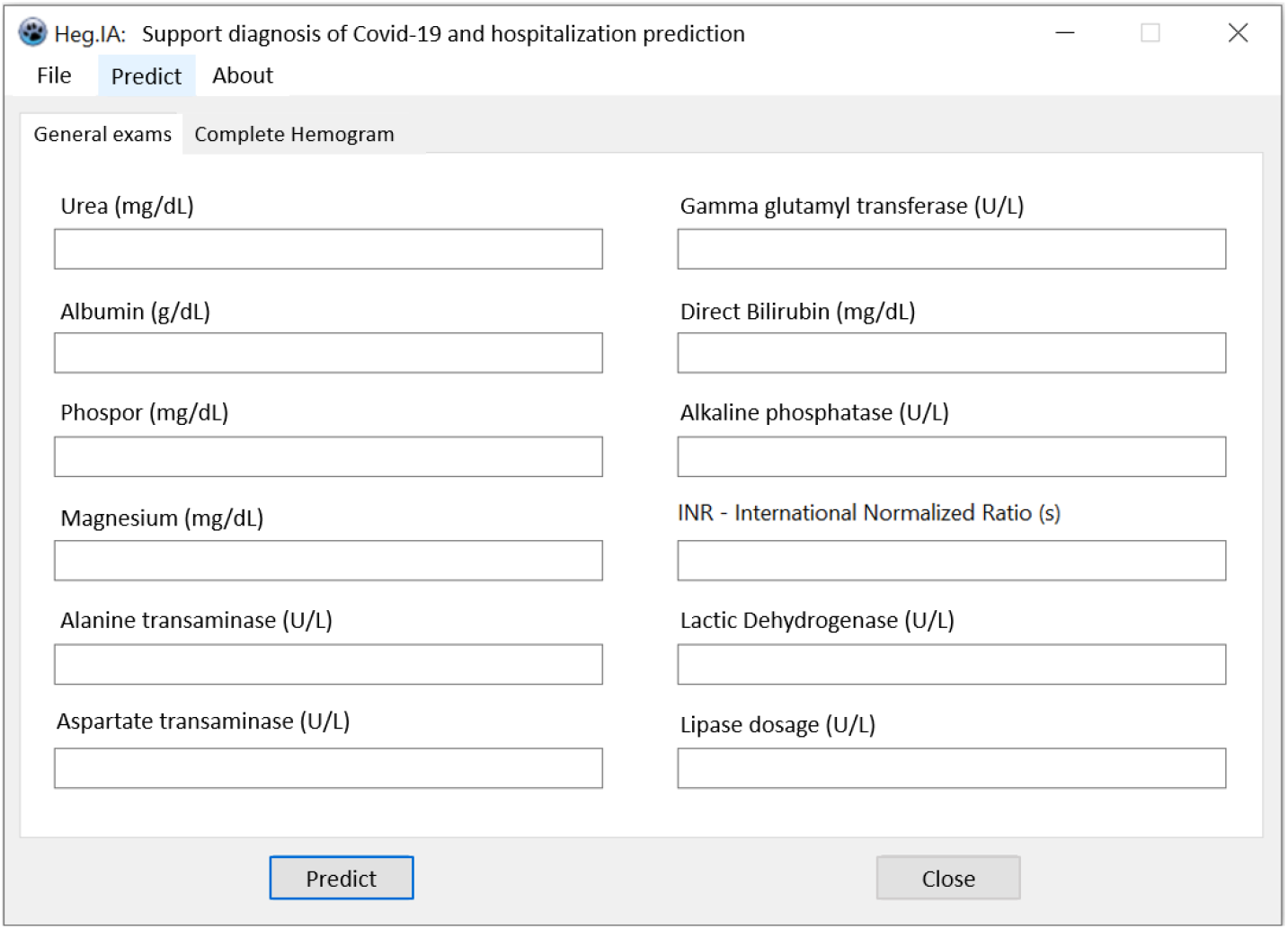
Screen 1 of the HegIA desktop application. General blood tests should be entered using the indicated units.

**Figure 7:**
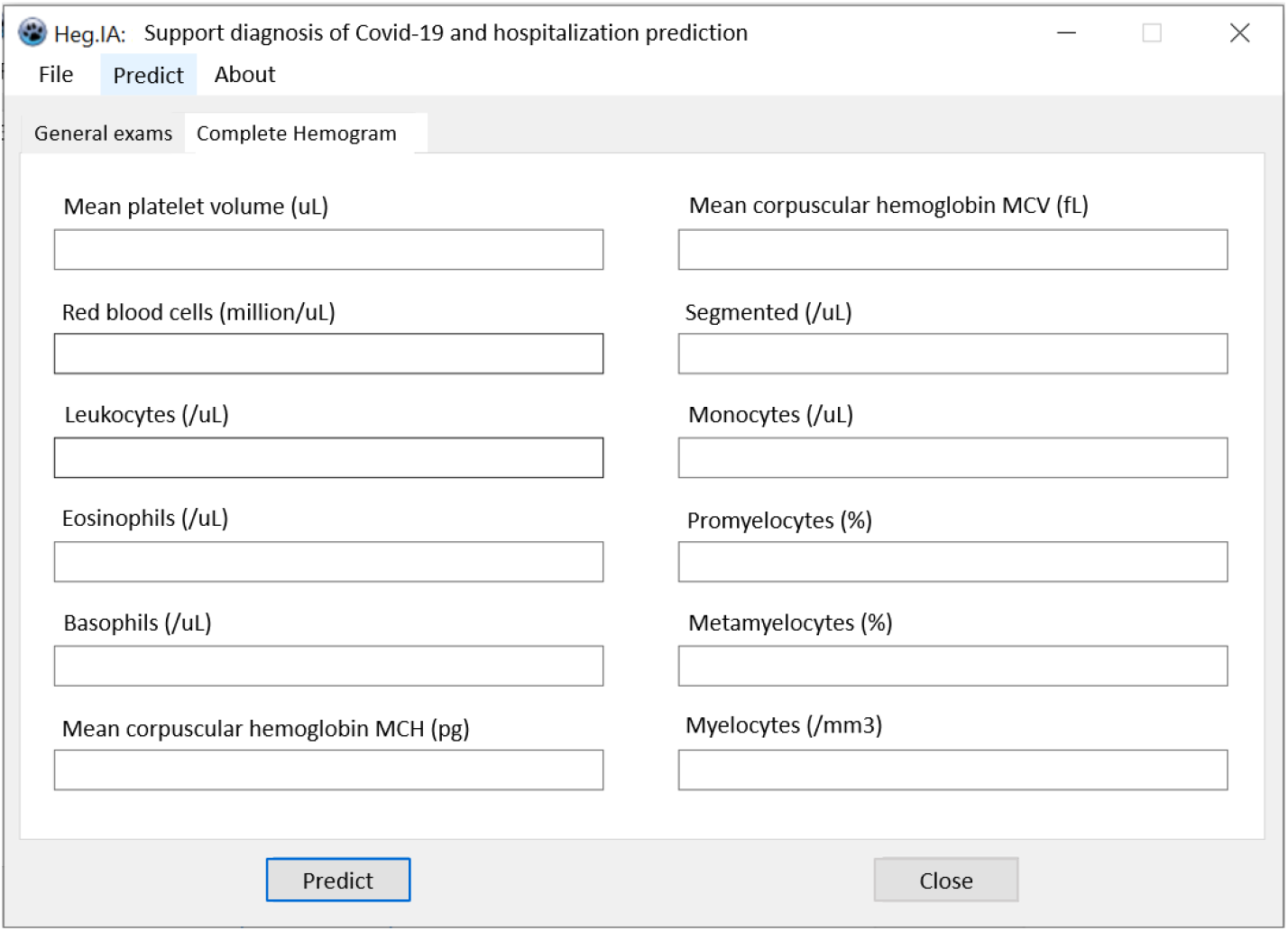
Screen 2 of the HegIA desktop application. Blood tests from complete hemogram should be inserted using the indicated units.

**Figure 8:**
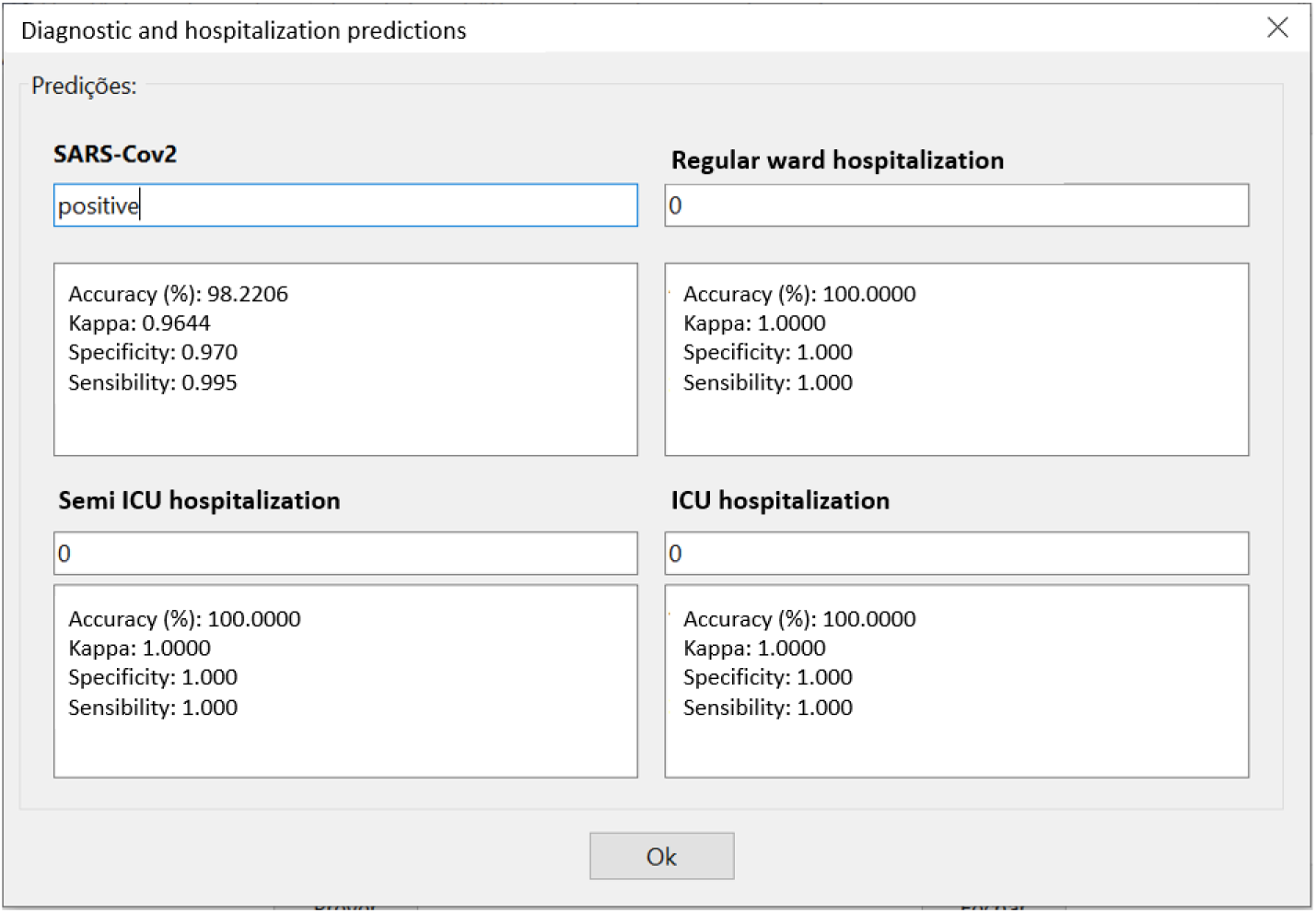
Covid-19 diagnostic prediction results screen from the HegIA desktop application.

Knowing that the classification method was trained with normalized database. Our desktop application rescales the data to fit with this data. The rescaling is made considering the reference values of each exams. So we could estimate each new value according the normalization.

The code for this desktop version is freely available for non-commercial purpose on Github repository: github.com/Biomedical-Computing-UFPE/Heg. IA-Desktop.

## 5. Discussion

The initial list of blood tests is quite extensive. In the context of researching a new disease, broader investigation is always needed. Thus, it is possible to verify the presence of other factors that may worsen the patient’s health status. Examples of this are altered glycemic indexes, chronic obstructive pulmonary diseases, systemic arterial hypertension, as well as infections by other groups of viruses. This investigation also makes it possible to assess the course of the disease in different groups of individuals. Despite the great relevance of this search, a group of 107 clinical examinations is very large in the context of a pandemic. They could lead to long periods of analysis. This highlights the need to apply methods to select the most important attributes.

In this way, during the feature selection phase our method was able to reduce the amount of features (exams) in more than 20%. This was an important achievement, since it reduces the amount of required procedures. Therefore resulting in less expensive and less time-consuming diagnostic process.

For the classification stage, Figures 4 and 5 show that, overall, Bayes Net method overcame the others in accuracy, which was greater than 95%, and in kappa statistic, reaching more than 0.90. All tested configurations of Random Forest also showed good performance, with accuracies around 95% and kappa close to 0.90. Naive Bayes algorithm performed slightly worse than these others, but still achieving great results for both accuracy and kappa. SVM, MLP and Random Tree classifiers, on the other hand, achieved less impressive results, with accuracies around 90% and kappa between 0.75 and 0.85. As for the data dispersion, it was slightly greater for kappa results, when compared to accuracy. However, both graphs show low dispersion, indicating good reliability of the system.

The superior results of the Bayes network may indicate or confirm the features (exams) are statistically independent or, at least, preserve a low level of interdependence. Therefore, even though all 24 selected exams showed to be relevant to diagnose SARS-Cov2, our hypothesis is that they could exhibit some degree of independence from each other.

The results presented in Table 3 show the classification performance using Bayes Net among the three databases: with the complete set of exams (original database), with features selected by PSO and the one including just the cheaper exams. The table shows the average and standard deviation values for all six metrics: accuracy, kappa statistic, recall, sensibility, precision and specificity. By comparing the results of all databases, we observed a minimal reduction in accuracy, recall and sensitivity with the decrease of the amount of features. All these metrics showed a decrease lower than 4%, when using only the cheap exams. For precision and specificity, the reduction for 24 exams led to a decrease of around 5%. The selection of these exams triggered a reduction of 8% in kappa statistic.

Thus, the 24 selected tests have been shown to be efficient in the diagnosis of Covid-19 in a fast way. Several studies have analyzed the clinical potential of these hematological parameters. Lippi & Plebani (2020) points out that many patients with Covid-19 do not have abnormal coagulation tests at the time of admission to the hospital. However, there is a gradual increase as the severity of the disease increases. Wan et al. (2020) also detected cases of hypercoagulable state and secondary hyperfibrinolysis. These findings confirm that parameters such as the mean volume of platelets are important in identifying severe cases and predicting the risk of mortality (Tang et al., 2020a; Lippi et al., 2020).

Lippi & Plebani (2020) also summarized the main contributions of laboratory tests to Covid-19. Among them are those of albumin, aspartate transaminase, alanine transaminase, bilirubin, lactate dehydrogenase and leukocytes. These same exams were selected by the methods in our work, confirming the relevance of the results obtained. The study of Huang et al. (2020) also showed cases of leukopenia and lymphopenia. This may indicate a decreased immunological response to the virus. On the other hand, the potential clinical significance of the increased value of aminotransferases and bilirubin aligned with low albumin values is a liver injury. Wan et al. (2020) suggest that these changes in the severe patients were more obvious. Liver damage might be directly caused by the viral infection of liver cells (Zhang et al., 2020a). High levels of lactate dehydrogenase can be related to pulmonary injury and/or widespread organ damage (Lippi & Plebani, 2020). In addition, Lippi & Plebani (2020) believes that increased monocytes levels can show a severe viral infection.

These results support the idea that Covid-19 can causes damages beyond hematological and respiratory issues. Examples are multi-organ failure (MOF) and its complications, and intravascular coagulopathy (Lippi et al., 2020). These results highlight the importance of diagnosis based on clinical examinations.

## 6. Conclusion

A new kind of coronavirus, SARS-Cov2, started the biggest pandemic of the century. This virus has a stronger human-to-human transmission capacity, and has already led to millions of infected people and thousands of deaths. One of the main strategies to fight the pandemic is testing in a precise and quick way (WHO, 2020).

The ground-truth test in Covid-19 diagnosis is the Reverse Transcription Polymerase Chain Reaction (RT-PCR) with DNA sequencing and identification. RT-PCR is precise, but takes several hours to be assessed. Another type of test, based on IgM/IgG antibodies, delivers results quickly, however they are nonspecific for Covid-19, and may have very low sensitivity and specificity. IgM/IgG tests do not directly detect the SARS-Cov2 presence, indeed they detect the serological evidence of recent infection.

Considering this, the development of a diagnosis support system to provide fast results with high sensitivity and specificity is necessary and urgent. In this context, blood tests have some advantages. First, they are commonly used during medical screening. Besides that blood tests are less expensive and less time-consuming than other diagnosis methods. Thus providing a more accessible system. By combining these blood tests results to analysis based on Artificial Intelligence, we were able to provide a robust, efficient and easily available system to diagnose Covid-19.

We optimized the system by reducing the amount of required exams, based on their relevance to describe the diagnosis problem and on their price and availability worldwide, specially in low-income communities. Firstly we performed an automatic exam selection using Particle Swarm Optimization (PSO) method. Then, we manually chose some exams aiming to reach an optimal combination of price, time and number of procedures. This procedure resulted in 24 blood tests, which can be delivered in up to one hour. Since the computational classification can be performed in milliseconds, with the 24 blood tests results, a technician can get diagnostic results relatively fast. The user just have to fill in the electronic form with these 24 blood tests results.

By using a computationally simple method, based on the classical Bayesian networks, we were able to achieve high diagnosis performance: 95.159% *±* 0.693 of overall accuracy, kappa index of 0.903 *±* 0.014, sensitivity of 0.968 *±* 0.007, precision of 0.938 *±* 0.010 and specificity of 0.936 *±* 0.011. If compared to deep-learning based methods, the proposed system also reduces computational cost.

More importantly, our approach provides a possible way out for the tests availability issue in the context of Covid-19. The availability of this software system combined to low cost and fast tests, based on blood analysis, may be of great help to overcome the testing challenges being experienced worldwide. Less-favored countries and communities can especially benefit from this solution. With this system we may be able to expand access to effective testing and thereby reach and save more people.

## Data Availability

The code is freely available at GitHub repository "Heg.IA-Desktop" at "Biomedical-Computing-UFPE".

https://github.com/Biomedical-Computing-UFPE/Heg.IA-Desktop

## Footnotes

* Wellington Pinheiro dos Santos Email address: (Wellington Pinheiro dos Santos)

## Acknowledgements

The authors are grateful to the Brazilian research agencies CAPES and CNPq, for the partial financial support of this research.

## Compliance with Ethical Standards

This study was funded by the Brazilian research agencies CAPES and CNPq.

All procedures performed in studies involving human participants were in accordance with the ethical standards of the institutional and/or national research committee and with the 1964 Helsinki declaration and its later amendments or comparable ethical standards.

## Footnotes

1 According to the Brazilian Society of Clinical Analysis, there is no reference document to establish standard turnaround times for the exams described above.

